# Screening for diabetes and impaired glucose metabolism in Qatar: models’ development and validation

**DOI:** 10.1101/2021.04.04.21254900

**Authors:** Khaled Sadek, Ibrahim Abdelhafez, Israa Al-Hashimi, Wadha Al-Shafi, Fatihah Tarmizi, Hissa Al-Marri, Nada Alzohari, Mohammad Balideh, Alison Carr

**Author notes:** **Correspondence:** Khaled Sadek; Ibrahim Abdelhafez, College of Medicine, Qatar University, 2713 Doha, Qatar. Both authors contributed equally to the manuscript (Khaled Sadek, Ibrahim Abdelhafez).

## Abstract

**Aim:** To establish two scoring models for identifying individuals at risk of developing Impaired Glucose Metabolism (IGM) and Type two Diabetes Mellitus (T2DM) in Qatari.

**Materials and Methods:** A sample of 2000 individuals, from Qatar BioBank, was evaluated to determine features predictive of T2DM and IGM. Another sample of 1000 participants was obtained for external validation of the models. Several scoring models screening for T2DM were evaluated and compared to the model proposed by this study.

**Results:** Age, gender, waist-to-hip-ratio, history of hypertension and hyperlipidemia, and levels of educational were statistically associated with the risk of T2DM and constituted the Qatari diabetes risk score (QDRISK). Along with, the 6 aforementioned variables, the IGM model showed that BMI was statistically significant. The QDRISK performed well with an area under the curve (AUC) 0.870 and 0.815 in the development and external validation cohort, respectively. The QDRISK showed overall better accuracy and calibration compared to other evaluated scores. The IGM model showed good accuracy and calibration, AUCs 0.796 vs. 0.774 in the development and external validation cohorts, respectively.

**Conclusions:** This study developed a Qatari-specific risk scores to identify high risk individuals and can guide the development of a nationwide primary prevention program.

## 1. Introduction

Diabetes is one of the most prevalent chronic diseases worldwide. The International Diabetes Federation (IDF) estimated that approximately 463 million individuals aged between 20 -79 are diagnosed with diabetes in 2019, and that this figure is expected to rise to 700 million by 2045 [1]. Qatar is reported as one of the most affected populations in the world with an age-adjusted prevalence of 15.6% [1]. Along with its high prevalence, diabetes contributes to a number of major complications including cardiovascular diseases, nephropathies, retinopathies, and neuropathies. [2]. Further, diabetes imposes a considerable economic burden, reported as $327.2 billion in the U.S in 2017 [3]

In addition, the most recent estimate of undiagnosed cases of diabetes was as high as 231.9 million. Consequently, early detection of such individuals might prevent or delay the onset of Type 2 Diabetes Mellitus (T2DM) as it contributes to 90% of diabetes [1]. Several risk factors were associated with increased incidence of T2DM, including; age, ethnicity, family history, obesity, nutrition, smoking, and physical inactivity [1, 2]. Screening for T2DM can be conducted with either non-invasive risk scores or invasive laboratory measure of glycosylated hemoglobin (HBA1c), fasting plasma glucose, or random blood glucose (RBG) levels.

In comparison to invasive laboratory testing, screening models are both time and cost effective, along with its convenient applicability in clinical settings [4]. Several screening risk scores for T2DM were developed for different populations [5-18]. To the best of our knowledge, there is no score developed screening for T2DM and Impaired Glucose Metabolism (IGM) using Qatari population data. Therefore, this study aims to generate two simple and reliable screening models, that is tailored specifically for the Qatari population and can be implemented within primary healthcare settings or by individuals in the community to identify ones at high risk of developing T2DM and IGM in Qatar.

## 2. Methods

### 2.1. Study design, setting and data source

This cross-sectional study was conducted based on data provided from the Qatar BioBank (QBB). QBB is a population-based project which recruits adults who are either nationals or long-term residents (≥ 15 years) in Qatar aged 18 to 89 years, and follows them up to measure a wide range of various health parameters. Recruitment of subjects started in 2012 and obtained information of more than 17,065 participants. Participation is voluntary, based on personal recommendations of friends and family, via social media, bookings via the website (available at: www.qatarbiobank.org.qa), or through phone calls.

Upon obtaining written informed consent from participants in their first visit to the QBB facility at Hamad Medical City Building 17, Doha, Qatar, they undergo a 5-stage interview, along with physical and clinical measurement sequence, the interview averaging 3 hours. Comprehensive self-reported questionnaires are completed on health behaviors, past medical history, lifestyle variables, physical activity, mental well-being, environmental factors’ exposure and other parameters, followed by physical examination to obtain anthropometric measurements, blood pressure, electrocardiogram, bone density and other measures by trained research personnel. Afterwards, enrolled participants provide saliva, blood and urine samples, which undergo analysis at Hamad General Hospital Medical Centre Laboratory in Doha, Qatar [19, 20].

Participants are contacted via e-mail, messages, or phone calls to present for a follow up visit at QBB facility every 5 years, where they undergo the same procedures as the baseline visit. QBB is regularly recruiting further participants, aiming to represent the population at Qatar by reaching 60,000 study participants. For detailed description of procedures of biological marker assessments please review Qatar Biobank for Medical Research [21]. Ethical approval for the study was obtained from the QBB Institutional Review Board (Ex-2018-RES-ACC-0097-0043).

### 2.2. Inclusion and exclusion criteria

A cross-sectional study was carried out involving participants provided by QBB aging 18 years or older. Candidates with a history of type 1 diabetes mellitus or gestational diabetes were excluded from this study. From the remaining participants, only those with HbA1c and random plasma glucose (RPG) levels available were included in the analysis.

### 2.3. Definitions of outcomes

#### 2.3.1. The Qatari diabetes risk score (QDRSIK)

Participants with HbA1c ≥6.5% and/or history of diabetes and/or history of anti-diabetic medication were considered diabetic individuals based on this proxy composite criterion. All other participants were deemed non-diabetic.

#### 2.3.2. Screening for impaired glucose metabolism

For comparative analysis of performance measures, we developed a second model screening for IGM, where participants with HbA1c level of ≥5.7% [2] and/or RPG levels ≥140 mg/dL (7.8 mmol) [22], and/or history of diabetes and/or history of anti-diabetic medication were deemed having IGM, while the remaining participants were labeled non-IGM.

### 2.4. Statistical analysis

Differences between categorical variables were compared using Chi-square test and Fisher’s Exact Test and were reported as frequencies and percentages, while differences between continuous variables was examined using Mann-Whitney test and were reported as median (IQR). To aid with clinical decision-making, continuous variables were dummy coded into (0 vs. 1) representing above and below cut-offs pre-determined. While this approach is often criticized, we believe it is practical and more applicable in primary healthcare settings.

### 2.5. Variable Selection and Score Construction

#### 2.5.1. Least Absolute Shrinkage and Selection Operator

To avoid over-fitting, ten variables including: age, gender, history of hypertension (HTN), history of hyperlipidemia (HLD), waist-to-hip ratio (≥1 for males and ≥0.86 for females), hours of sleep, BMI (normal, overweight, and obese) [23], educational level, vegetable and fruit consumption were entered into the Least Absolute Shrinkage and Selection Operator (LASSO) binary logistic regression against the two composite binary outcomes detailed above.

The L1-penalized LASSO was utilized for the multivariable analysis with 10-fold cross validation for internal validation. LASSO is a machine-learning logistic regression which penalizes the size of the coefficients of based on the value of the hyperparameter lambda (λ). Coefficients of weaker variables are reduced towards zero with larger penalties, while only the strongest features remain in the model. Feature selection was carried out by the minimum value of lambda (λ.min). The R package “glmnet” was used to conduct the LASSO regression. Afterwards, variables selected by LASSO entered a logistic regression model.

#### 2.5.2. Complex Machine Learning Analysis

The second strategy used in this study one relied on further sophisticated machine learning (ML) analysis using the final significant variables in the logistic regression model to build four ML models, using random forest (RF), gradient boosting machine (GBM), XgBoost as well as deep learning (DL). These models also used 10-fold cross-validation. Statistical analysis for the aforementioned ML models was performed using R package “h2o” (version 3.32.1.1).

### 2.6. External validation of several diabetes risk scores

We attempted to externally validate diabetes risk score where data for their categories was available. The choice of these models was either based on practicality and ease of use. These included the Danish [24], Thai [17], and the Data from the Epidemiological Study on the Insulin Resistance Syndrome (DESIR) scores [25]. Further, we tested the hypothesis that including models developed in populations with similar characteristics in the gulf region would yield the maximum accuracy and calibration measures. Therefore, we assessed the performance of models developed in the United Arab Emirates (UAE) [26], Oman [27], along with the Qatari mathematical model generated using data simulation techniques [28].

### 2.7. Assessment of accuracy and calibration

The accuracy of QDRSIK was assessed using the area under the receiver-operator characteristic curve (AUC) of the logistic and ML models and compared them to those of the six aforementioned-mentioned models. Calibration of the models was assessed using the “rms” package in R with 200 bootstrapped resamples of the AUC, McKelvoy’s R^2^, Brier score, calibration slope, intercept values. In addition, we used the Hosmer–Lemeshow goodness of fit test (HL) χ^2^ value with <20 and p-values ≥0.01 indicating adequate calibration [29]. All statistical analyses were performed with R software (version 4.0.4, R Foundation), and statistical significance was attained at *p-values*<.05.

## 3. Results

### 3.1. Participant’s characteristics

A random sample of 2000 participants was obtained from QBB for the model development (2018). Data of 1000 individuals was obtained from QBB at a later timepoint (2021) for external validation. After excluding duplicates along with participants with missing data in variables defining the outcome and/or other key variables in both datasets, only 1660 and 930 participants remained in the model development and the validation cohorts, respectively, and underwent further analysis. The male gender comprised 895 participants (53.9%) in the development cohort with a a median age of 37 (IQR: 30-47.2) years, and a median BMI of 28.29(IQR: 25.1-32.3) kg/m2. In addition, the median HbA1c was 5.40% (IQR: 5.10-5.80), while RPG was 5.10 (IQR: 4.74-5.6) mmol. Participants were grouped into diabetic (n = 169, 10.2%) and non-diabetic (n = 1491, 89.8%). Compared with the Non-diabetics, patients with T2DM participants were more likely to be older, obese, with history of HTN and HLD, and with lower educational levels (Table 1). In the second model, participants were categorized into those with IGM (n = 606, 32.6%) and non-IGM (n = 1254, 67,4%). Those with IGM were more likely to be older, obese, with history of HTN and HLD (Table 1). Subjects in the external validation cohort had similar overall characteristics compared to the development cohort (Table 2).

**Table 1:**
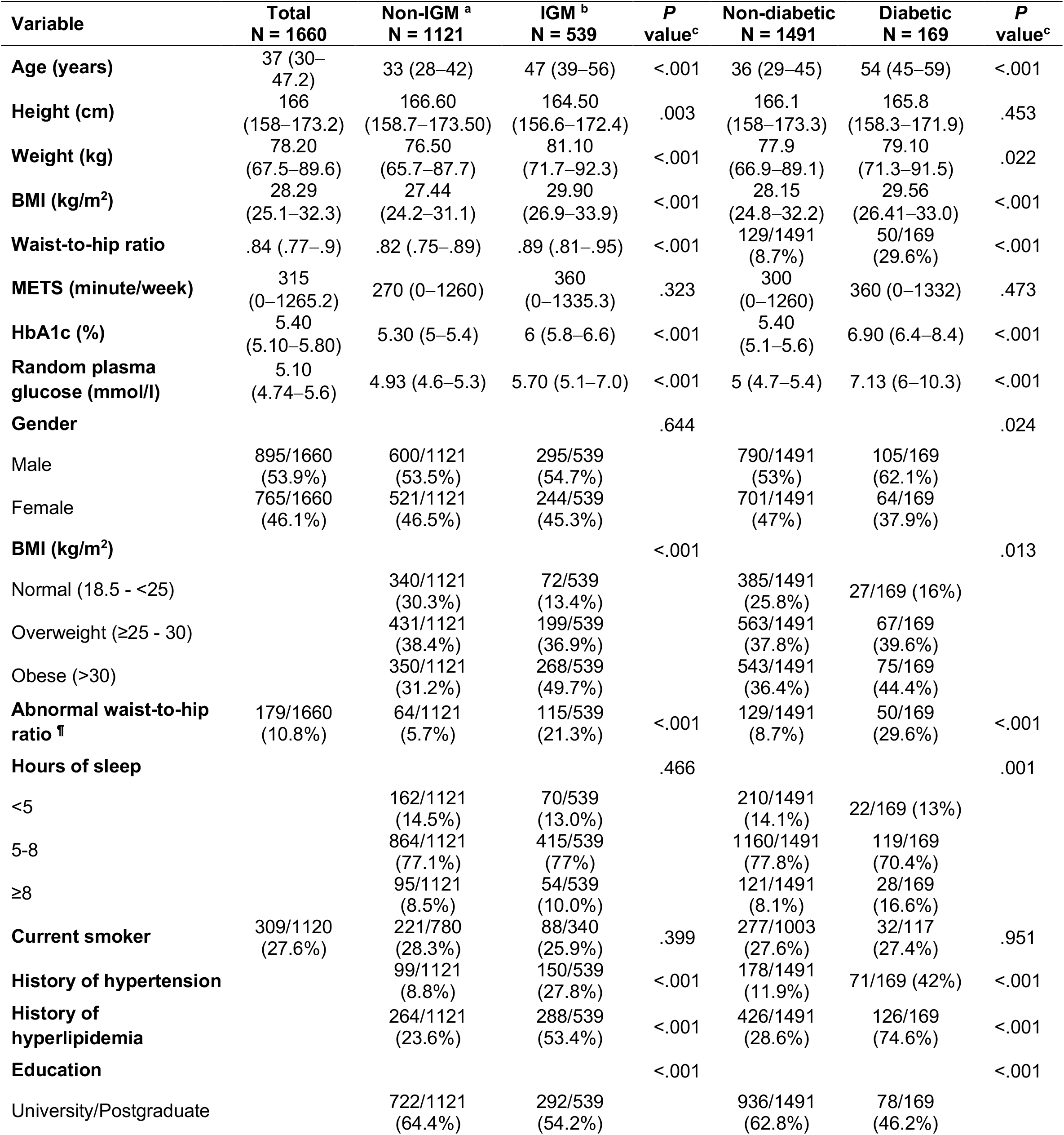
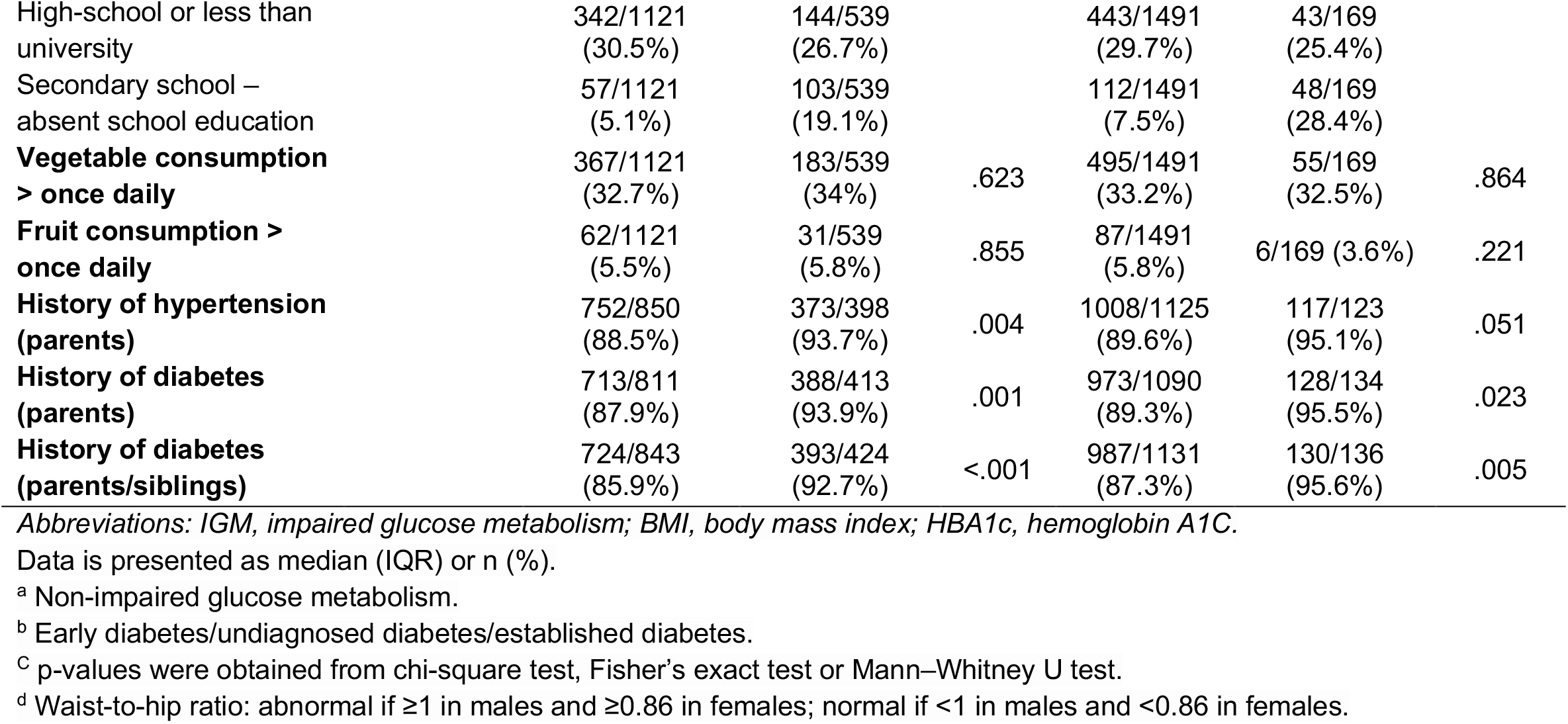
Characteristics of the model development cohort based on diabetic status and impaired glucose metabolism.

**Table 2:**
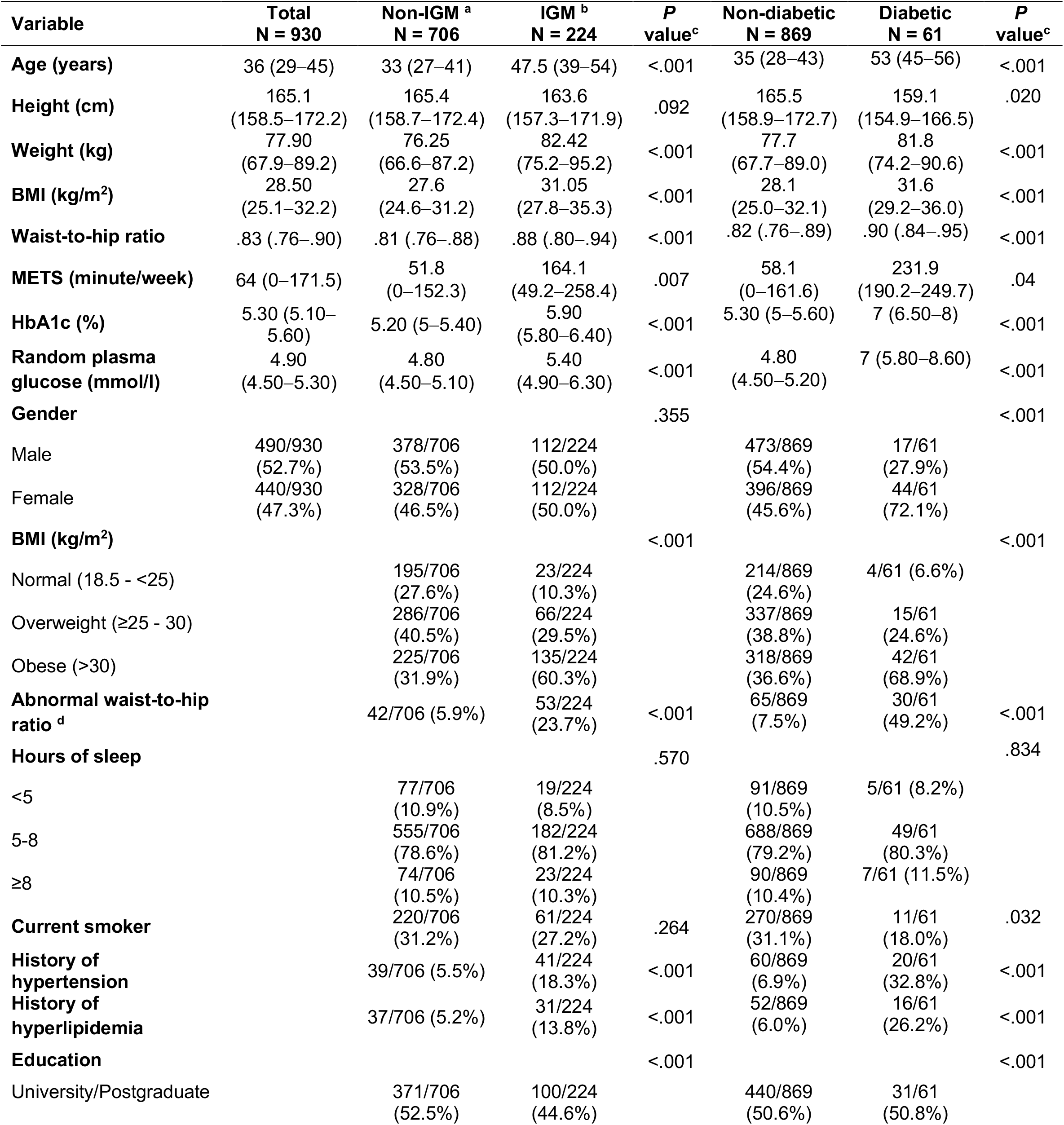
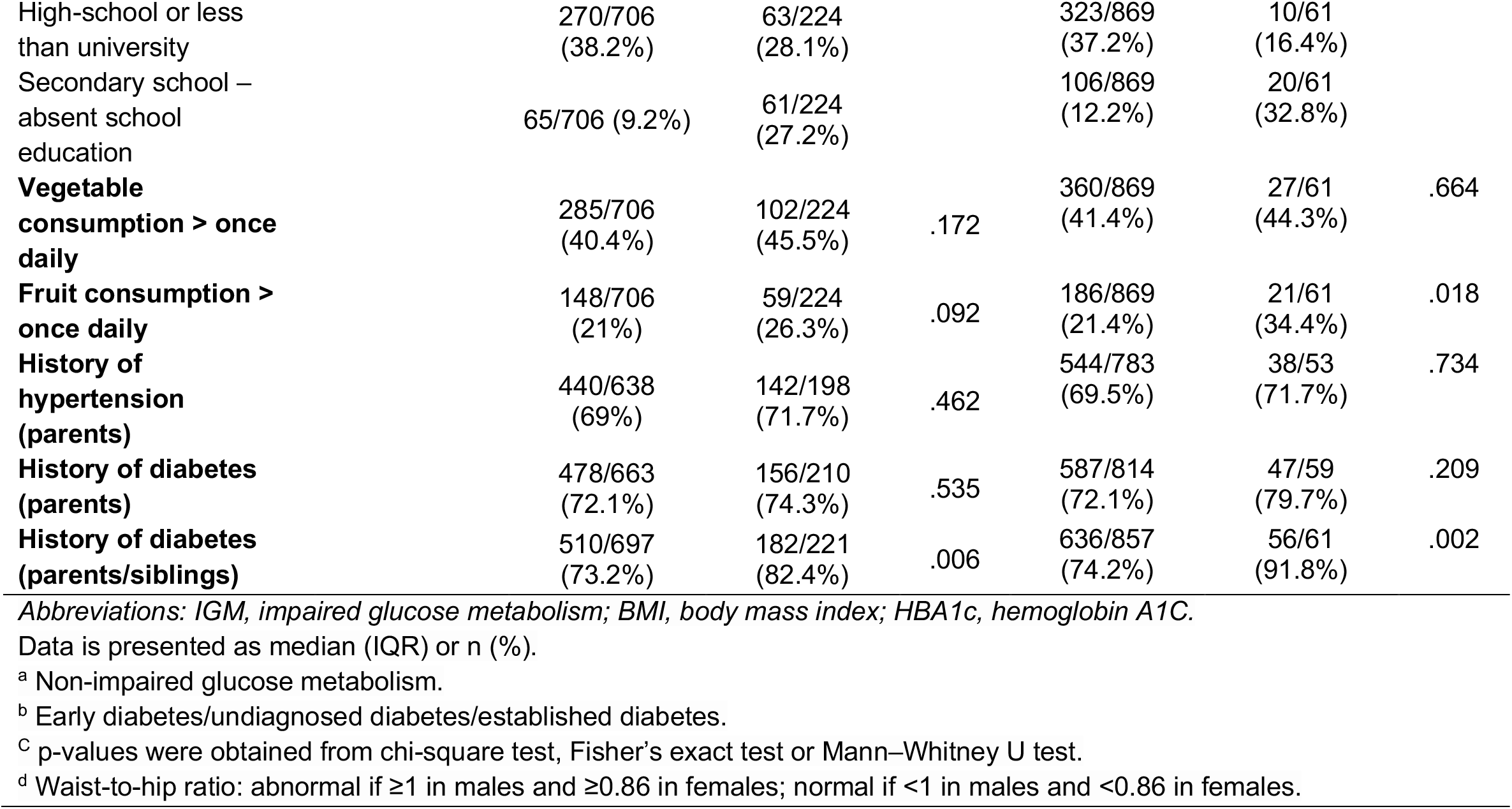
Characteristics of the external validation cohort based on diabetic status and impaired glucose metabolism.

**Table 3:** Multivariable logistic regression and assigned scores for variables predicting Type 2 Diabetes Mellitus (T2DM).

| Variable | $\beta$ coefficient | P value | OR (95% CI) | Risk score |
| --- | --- | --- | --- | --- |
| **Age (years)** |  |  |  |  |
| 18–36 | Reference |  |  | 0 |
| 36–54 | 1.21 | <.001 | 3.35 (1.89–6.28) | 12 |
| ≥55 | 2.62 | <.001 | 13.83 (7.39–27.21) | 26 |
| **Gender** |  |  |  |  |
| Female | Reference |  |  | 0 |
| Male | .87 | <.001 | 2.40 (1.61–3.65) | 9 |
| **Waist-to-hip-ratio** |  |  |  |  |
| Normal | Reference |  |  | 0 |
| Abnormal | .65 | .005 | 1.93 (1.20–3.05) | 7 |
| **History of hyperlipidemia (yes)** | 1.19 | <.001 | 3.30 (2.20–4.99) | 12 |
| **History of hypertension (yes)** | .70 | .002 | 2.02 (1.34–3.01) | 7 |
| **Education** |  |  |  |  |
| University/Postgraduate | Reference |  |  | 0 |
| High-school or less than university | .16 | .478 | 1.17 (.74–1.82) | 2 |
| Secondary school – absent school education | .76 | .003 | 2.14 (1.27–3.55) | 8 |
| **Hours of sleep** |  |  |  |  |
| <5 | Reference |  |  |  |
| 5–8 | -.27 | .34 | .75 (.43–1.37) | 0 |
| ≥8 | .60 | .10 | 1.82 (.89–3.76) | 0 |
| Vegetable consumption (< once daily) | - | - | - | 1 |
| Fruit consumption (< once daily) | - | - | - | 1 |
| **Cutoff point <sup>†</sup>** |  |  |  | **31** |

Abbreviations: OR, odds ratio; CI, confidence interval.
<sup>†</sup> The cut-off point was obtained from the optimal Youden index.
Scores 0-31, 32-42, >42 were deemed as low, moderate, and high risk of developing T2DM, respectively.

### 3.2. The Qatari diabetes risk score (QDRISK)

#### 3.2.1. Logistic regression

The LASSO analysis excluded the following variables: BMI, vegetable and fruit consumption (Figures S1-A & S1-B). Subsequently, the remaining 7 variables (age, gender, waist-to-hip-ratio, history of HTN, history of HLD, educational levels and hours of sleep) entered a logistic regression (LR) analysis and 6 of them were found to be significant predictors of T2DM. Beta-coefficients of significant variables were multiplied by 10 and rounded to the nearest integer and therefore constructed the risk score. These included age ≥55 (26 points), 36–54 (12 points), history of HLD (12 points), male gender (9 points), abnormal Waist-to-hip-ratio (7 points), history of HTN (7 points), education levels: high-school or less than university (2 points), secondary school or absent school education (8 points) (please see Table 1). Although vegetable and fruit consumption (< once daily) were excluded by the LASSO, they were allocated 1 point each to highlight the role of lifestyle changes in prevention of T2DM [30-32].

The performance of the LR model in the training sample was tested by constructing a ROC curve. The AUC of the LR model in the development cohort was found to be 0.870 (95%CI 0.843–0.896) (Figure 1-A) compared to 0.815 (95% CI: 0.765-0.864) in the external validation cohort (Figure 1-B). The risk score cut-off (31 points) was obtained from the optimal Youden index with 82.2% sensitivity, 79.4% specificity, 31.2% positive predictive value (PPV) and 97.53 negative predictive value (NPV). The remaining values for the Qatari diabetes risk score (QDRISK) in the development and the validation sets are found in Table S1. Accordingly, subjects with 0-31, 32-42, >47 scores were deemed as at low, moderate, and high risk of developing T2DM, respectively. The model showed overall good calibration curves in both cohorts (Figure S2).

**Figure 1.**
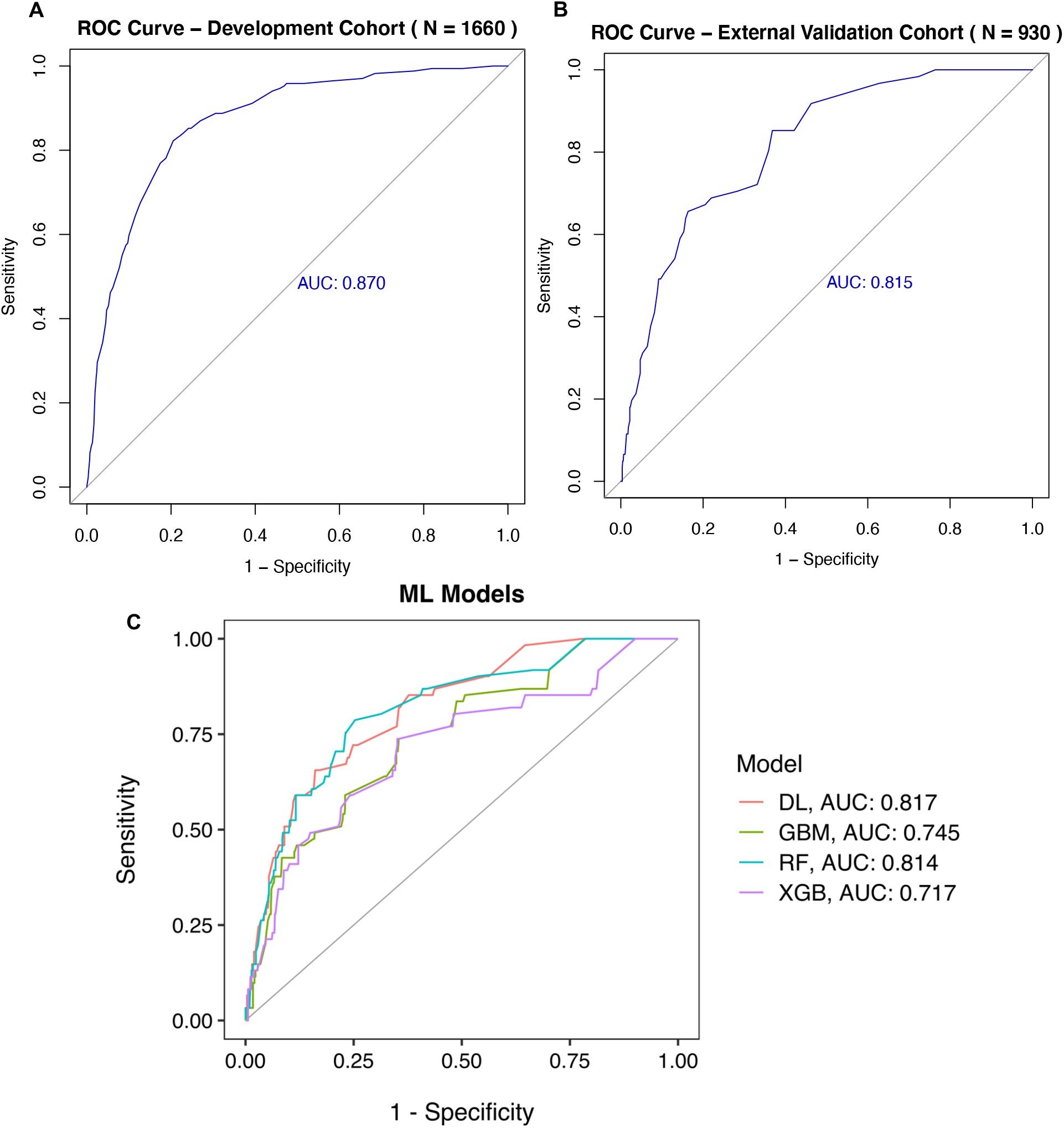
ROC curve showing the performance of the developed risk score in predicting T2DM with AUC, 0.870 (95%CI 0.843–0.896) in the development cohort **(A)** and 0.815 (95% CI: 0.765-0.864) in the external validation cohort **(B)**. ROC curve for predicting T2DM using machine learning models. Deep learning (DL), gradient boosting machine (GBM), random forest (RF), XgBoost (XGB) **(C)**.

#### 3.2.2. Comparative analysis with complex machine learning models

The performance of the ML models in the validation cohort was comparable to that of the regression model (Figure 1-A & 1-B) with DL AUC 0.817, GBM AUC 0.7452, RF AUC 0.814, and XGB AUC 0.717 (Figure 1-C). Sensitivities, specificities, PPVs and NPVs can be found in Table S2.

### 3.3. Screening for impaired glucose metabolism

The LASSO feature-selection excluded only vegetable and fruit consumption (Figure S1-C & S1-D), with the remaining 8 variables entered the LR analysis and remained significant predictors of IGM status. We also added vegetable and fruit consumption to the model despite being excluded by LASSO for the reasons highlighted earlier. Variables predictive of IGM and their scores can be found in Table S3. The discriminative ability of IGM score was lower compared to the QDRISK with AUC, 0.796 (95% CI: 0.774-0.819) in the development cohort (Figure S3-A) and 0.774 (95% CI: 0.740-0.809) in the external validation cohort (Figure S3-B) with overall good calibration (Figure S4). Performance of the ML models predicting IGM can be found in Figure S3-C.

### 3.4. External validation of diabetes risk scores

We were able to calculate diabetes risk scores in 1154, 2185, 2097, 2185, 1800, 976, and 1344 subjects for the Danish [24], UAE [26], Omani [27], Thai [17], Qatari mathematical model [28], as well as the female and the male versions of the DESIR models [25], respectively. Scores allocated to variables used in the aforementioned models are shown in Table S4. Despite hypothesizing higher performance for the UAE [26] and Oman [27] that were developed in the region, along with and Qatari mathematical model [28]; generated using the Qatari population characteristics and estimates of various disease prevalence; they all showed lower performance compared to the QDRISK with AUCs ranging (0.733-0.796) (Figure 2) and comparable calibration curves and other validation metrics (Figure S5 and Table S5). Interestingly, performance of the Danish score [24] showed better predictive accuracy in detecting T2DM compared to the QDRISK with AUC 0.824 (95%CI, 0.778-0.8694) (Figure 1-G), however, the overall lower validation metrics, with HL goodness of fit test (χ^2^ = 13.65, df: 8, p-value: 0.091) vs. (χ^2^ = 4.78, df: 8, p-value: 0.780). Of note, the DESIR models for females and males had the lowest predictive accuracy with AUCs 0.555 (0.508-0.635) vs. 0.486 (0.429-0.554) (Figure 1-A & 1-B), respectively, along with inferior calibration curves compared to the rest of the models. Therefore, we did not assess their validation metrics. Comparison of validation of the QDRSIK and externally validated models is shown in Table S5.

**Figure 2:**
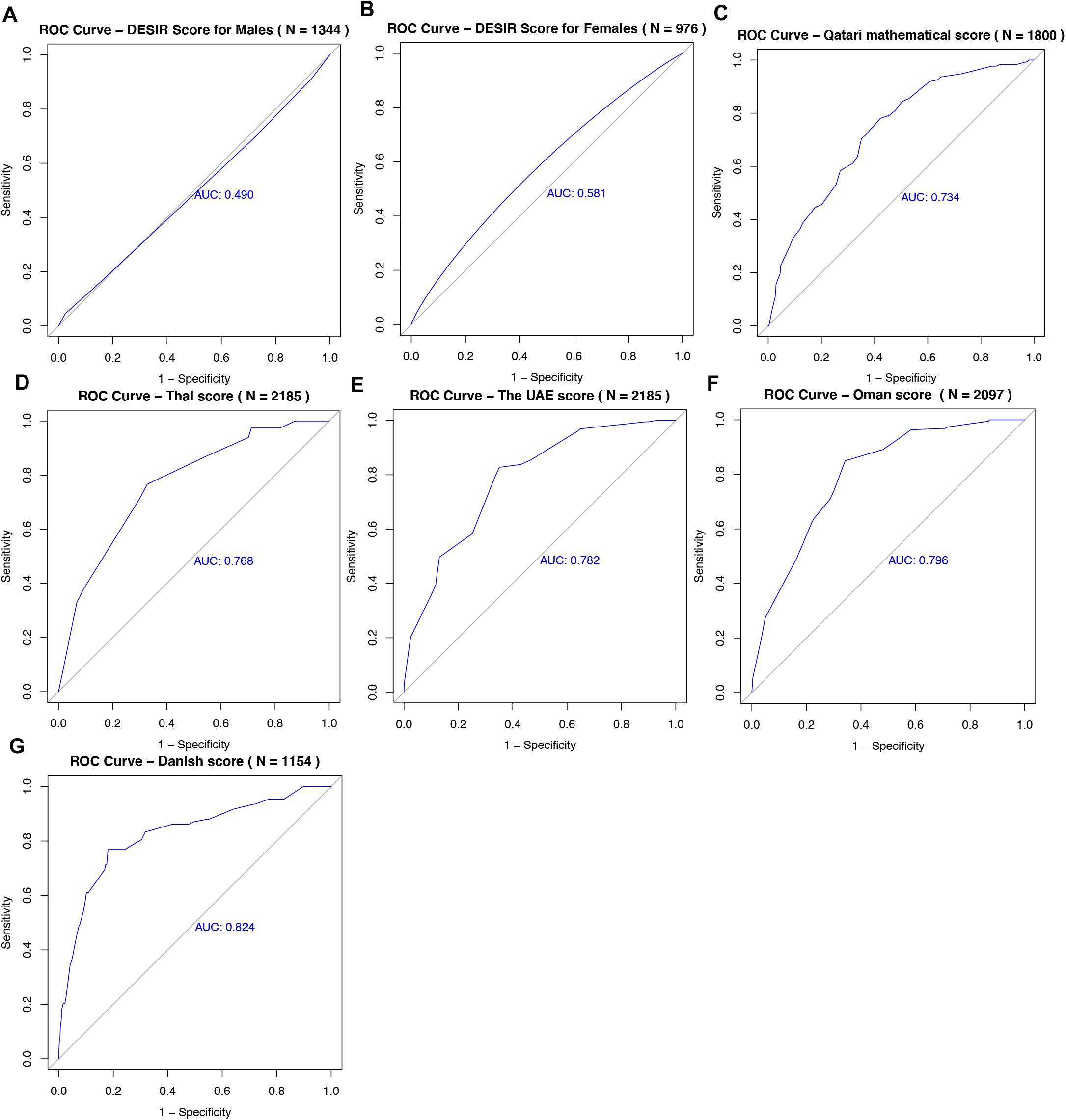
ROC curves showing comparable performance of the externally validated risk scores, with AUCs ranging (0.734-0.824) except for the DESIR scores (A & B).

## 4. Discussion

This study presents the first screening tool tailored specifically for the Qatari population, that identifies individuals at high risk of developing T2DM. The developed tool is based on 8 risk variables including; age, gender, waist-to-hip-ratio, history of HTN, history of HLD, educational levels, along with vegetable and fruit consumption. The proposed score can be utilized by healthcare professionals, or individuals in the community as the risk factors are easy to obtain and measure.

Age, gender, waist-to-hip-ratio, history of HTN, history of HLD, educational levels, along with vegetable and fruit consumption were identified as predictors for the risk of developing T2DM in previous risk scores [5-18, 28]. Most of these models had a reasonable performance in predicting the risk of T2DM in certain populations, with AUCs values ranging (0.52 -0.87). These results are similar to the model established in this study, with reasonable performance (AUC, 0.81).

Age was the highest predictive variable for the risk of T2DM in our score. This finding is consistent with other established scores [6, 7, 9, 10, 14, 18]. However, age was non-predictive in the score by Wilson et al. [12]. Several studies proposed that the risk of developing T2DM increases with older ages [6-10, 13-15, 33]. The male gender was also an important factor in our model in parallel with the studies by Aekplakorn, Wang and Kahn et al. [13, 15, 17].

Our study was not the first to include history of HTN and HLD in the risk score, as HTN showed significant prediction of the risk of T2DM in previous scores [6-10, 12-17], along with HLD [11-13, 15, 16]. However, HTN was not a predictor in the score by Chien et al. [11]. In addition, obesity significantly contributed to the established score as well as others [6-13, 16-18].

Based on the developed risk score values, (74.3% vs. 87.7%) of our sample were at low risk of developing T2DM compared to (15.3% vs. 7.4%) at moderate, and (10.1% vs. 4.8%) at high risk in the development and validation cohorts, respectively. In addition, (69.3% vs. 79.9%) of our sample were at low risk of having IGM compared to (19.6% vs. 13.7%) at moderate, and (11.1% vs. 6.5%) at high risk in the development and validation cohorts, respectively. Hence the need for screening at the level of the population and downstream intervention programs to target this proportion. Several diabetes prevention programs have been developed, including the Diabetes Prevention Program (DPP) [32] and the study by Tuomilehto et al [34]. Over three years, lifestyle interventions reduced the incidence of T2DM by about 60% in both studies [32, 34]. Thus, primary prevention of T2DM can be achieved through lifestyle interventions.

Compared to an estimated $77,445 10-year medical expenditures attributed to a patient with diabetes [3, 35], the 10-year costs of lifestyle interventions per capita were $4,601 in the DPP and its Outcome study [36]. This highlights the substantial cost-saving capability of lifestyle changes. Therefore, high risk subjects should be tested for T2DM, and implementation of lifestyle interventions for those with negative results along with moderate-risk subjects (13-27).

Notwithstanding its potential public health implications, two limitations to this study are important to highlight. First, we could not perform multiple imputation on family history of diabetes and HTN due to missing data (> 20%). Lack of this variable in our score might be a limitation, since it reflects the genetic predisposition for diabetes and is a significant factor that could increase the risk for T2DM [37, 38]. Nonetheless, it was not included in the widely implemented Finnish risk score, and the accurate German score [5, 8]. Finally, the use of cohort design could have improved the generalizability and reliability of the findings. On the other hand, our study was specifically designed and validated for the Qatari population, along with including an adequate sample size with a wide age range. The key advantages of this tool are its cost-effectiveness, practicality, and relying on factors that are not difficult to obtain. Hence, we recommend applying it at the level of the population as a screening tool for identification of high-risk individuals who would benefit from lifestyle interventions. Further studies should externally validate this risk score in larger cohorts within the Qatari population in the future.

In conclusion, our study investigated risk factors and developed a validated risk score designed for the Qatari population to assess the risk of developing T2DM as well as IGM based on demographic and anthropometric factors. It demonstrates a simple, cost-effective and convenient tool which may be used to identify individuals at high risk. Such individuals should undergo further blood testing and early lifestyle modifications aiming to decrease the incidence of T2DM and IGM. Further studies are needed to validate the utility of the present risk scores in larger cohorts in Qatar and externally.

## Data Availability

Although the corresponding authors Khaled Sadeq; and Ibrahim Abdelhafez, 2713 Doha, Qatar; wish to make data regarding this manuscript available online or through request, we declare that it cannot be made available under the policies of QBB related to data-sharing and participants' confidentiality.

## Author Contributions

Conceptualization, K.W.S.; I.A. and I.A.A.; methodology, K.W.S. and I.A.; validation, K.W.S. and I.A.; formal analysis, K.W.S. and I.A.; investigation, I.A.A., H.H.A. and N.A.; data curation, K.W.S. and I.A.; writing—original draft preparation, K.W.S., I.A. and I.A.A.; writing—review and editing, K.W.S., I.A., I.A.A., W.S.A., F.A.A., H.H.A., N.A. and M.A.B.; supervision, A.C. All authors have read and agreed to the published version of the manuscript.

## Acknowledgment

The publication of this article was funded by the Qatar National Library. We would like to thank Qatar Biobank project for providing the data.

## References

1. Federation, I.D., IDF Diabetes Atlas, 9th edn. 2019: Brussels, Belgium.

2. American Diabetes, A., Diagnosis and classification of diabetes mellitus. Diabetes Care, 2014. 37 Suppl 1: p. S81–90.

3. American Diabetes, A., Economic Costs of Diabetes in the U.S. in 2017. Diabetes Care, 2018. 41(5): p. 917–928.

4. Echouffo-Tcheugui, J.B., et al., Screening for type 2 diabetes and dysglycemia. Epidemiol Rev, 2011. 33(1): p. 63–87.

5. Schulze, M.B., et al., An Accurate Risk Score Based on Anthropometric, Dietary, and Lifestyle Factors to Predict the Development of Type 2 Diabetes. Diabetes Care, 2007. 30(3): p. 510–515.

6. Chen, L., et al., AUSDRISK: an Australian Type 2 Diabetes Risk Assessment Tool based on demographic, lifestyle and simple anthropometric measures. Med J Aust, 2010. 192(4): p. 197–202.

7. Glümer, C., et al., A Danish Diabetes Risk Score for Targeted Screening. The Inter99 study, 2004. 27(3): p. 727–733.

8. Lindstrom, J. and J. Tuomilehto, The Diabetes Risk Score: A practical tool to predict type 2 diabetes risk. Diabetes Care, 2003. 26(3): p. 725–731.

9. Al-Lawati, J.A. and J. Tuomilehto, Diabetes risk score in Oman: A tool to identify prevalent type 2 diabetes among Arabs of the Middle East. Diabetes Research and Clinical Practice, 2007. 77(3): p. 438–444.

10. Sulaiman, N., et al., Diabetes risk score in the United Arab Emirates: a screening tool for the early detection of type 2 diabetes mellitus. BMJ Open Diabetes Research & Care, 2018. 6(1): p. e000489.

11. Chien, K., et al., A prediction model for type 2 diabetes risk among Chinese people. Diabetologia, 2008. 52(3): p. 443.

12. Wilson, P.F., et al., Prediction of incident diabetes mellitus in middle-aged adults: The framingham offspring study. Archives of Internal Medicine, 2007. 167(10): p. 1068–1074.

13. Wang, A., et al., Risk scores for predicting incidence of type 2 diabetes in the Chinese population: the Kailuan prospective study. Scientific Reports, 2016. 6: p. 26548.

14. Khalaf, M., et al., Screening for diabetes in Kuwait and evaluation of risk scores. Vol. 16. 2010. 725–31.

15. Kahn, H.S., et al., TWo risk-scoring systems for predicting incident diabetes mellitus in u.s. adults age 45 to 64 years. Annals of Internal Medicine, 2009. 150(11): p. 741–751.

16. Balkau, B., et al., Predicting diabetes: clinical, biological, and genetic approaches: data from the Epidemiological Study on the Insulin Resistance Syndrome (DESIR). Diabetes Care, 2008. 31(10): p. 2056–61.

17. Aekplakorn, W., et al., A risk score for predicting incident diabetes in the Thai population. Diabetes Care, 2006. 29(8): p. 1872–7.

18. Mohan, V., et al., A simplified Indian Diabetes Risk Score for screening for undiagnosed diabetic subjects. J Assoc Physicians India, 2005. 53: p. 759–63.

19. Al Kuwari, H., et al., The Qatar Biobank: background and methods. BMC Public Health, 2015. 15(1): p. 1208.

20. Al Thani, A., et al., Qatar Biobank Cohort Study: Study Design and First Results. Am J Epidemiol, 2019. 188(8): p. 1420–1433.

21. Qatar Biobank for Medical Research. January 20, 2021]; Available from: http://www.qatarbiobank.org.qa/home

22. Engelgau, M.M., et al., Screening for diabetes mellitus in adults. The utility of random capillary blood glucose measurements. Diabetes Care, 1995. 18(4): p. 463–6.

23. Organization., W.H., Obesity and Overweight Fact Sheet. 2017.

24. Glümer, C., et al., A Danish diabetes risk score for targeted screening: the Inter99 study. Diabetes Care, 2004. 27(3): p. 727–33.

25. Balkau, B., et al., Predicting diabetes: clinical, biological, and genetic approaches: data from the Epidemiological Study on the Insulin Resistance Syndrome (DESIR). Diabetes Care, 2008. 31(10): p. 2056–61.

26. Sulaiman, N., et al., Diabetes risk score in the United Arab Emirates: a screening tool for the early detection of type 2 diabetes mellitus. BMJ Open Diabetes Res Care, 2018. 6(1): p. e000489.

27. Al-Lawati, J.A. and J. Tuomilehto, Diabetes risk score in Oman: a tool to identify prevalent type 2 diabetes among Arabs of the Middle East. Diabetes Res Clin Pract, 2007. 77(3): p. 438–44.

28. Awad, S.F., et al., A diabetes risk score for Qatar utilizing a novel mathematical modeling approach to identify individuals at high risk for diabetes. Sci Rep, 2021. 11(1): p. 1811.

29. Hosmer, D.W. and N.L. Hjort, Goodness-of-fit processes for logistic regression: simulation results. Stat Med, 2002. 21(18): p. 2723–38.

30. Tuomilehto, J., et al., Prevention of Type 2 Diabetes Mellitus by Changes in Lifestyle among Subjects with Impaired Glucose Tolerance. New England Journal of Medicine, 2001. 344(18): p. 1343–1350.

31. Pan, X.R., et al., Effects of diet and exercise in preventing NIDDM in people with impaired glucose tolerance. The Da Qing IGT and Diabetes Study. Diabetes Care, 1997. 20(4): p. 537–44.

32. Knowler, W.C., et al., Reduction in the incidence of type 2 diabetes with lifestyle intervention or metformin. N Engl J Med, 2002. 346(6): p. 393–403.

33. Halter, J.B., Aging and Insulin Secretion, in Handbook of the Biology of Aging, S.N.A. Edward J. Masoro, Editor. 2011, Elsevier Inc. p. 373–384.

34. Tuomilehto, J., et al., Prevention of type 2 diabetes mellitus by changes in lifestyle among subjects with impaired glucose tolerance. N Engl J Med, 2001. 344(18): p. 1343–50.

35. American Diabetes, A., Economic costs of diabetes in the U.S. in 2012. Diabetes Care, 2013. 36(4): p. 1033–46.

36. Diabetes Prevention Program Research, G., The 10-year cost-effectiveness of lifestyle intervention or metformin for diabetes prevention: an intent-to-treat analysis of the DPP/DPPOS. Diabetes Care, 2012. 35(4): p. 723–30.

37. Rich, S.S., Mapping Genes in Diabetes: Genetic Epidemiological Perspective. Diabetes, 1990. 39(11): p. 1315–1319.

38. Kekalainen, P., et al., Hyperinsulinemia cluster predicts the development of type 2 diabetes independently of family history of diabetes. Diabetes Care, 1999. 22(1): p. 86–92.

