## Supplementary for "Screening for diabetes and impaired glucose metabolism in Qatar: models’ development and validation"

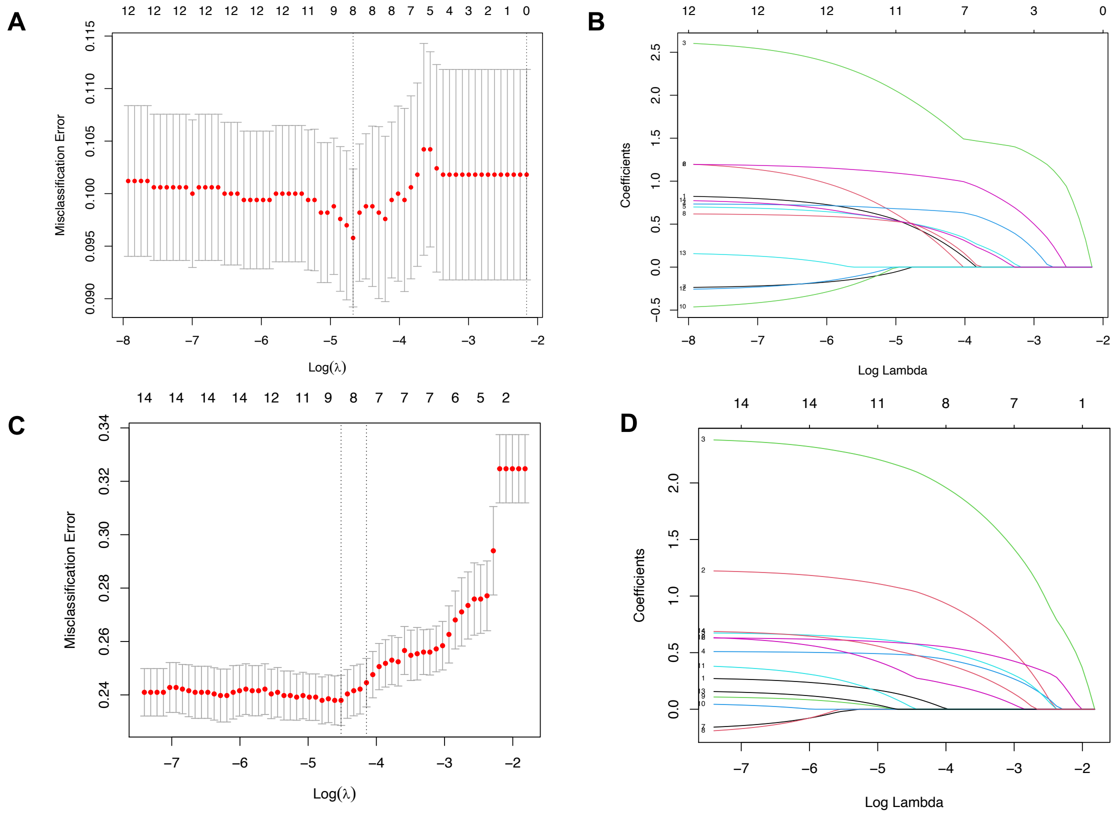


**Figure S1.** Variable selection using the least absolute shrinkage and selection operator (LASSO) logistic regression model. The tuning parameter lambda (λ) selection in the LASSO model via 10-fold cross-validation using the minimum value (A: Diabetes, C: IGM). LASSO coefficient profiles of the 10 variables (B: Diabetes, D: IGM).

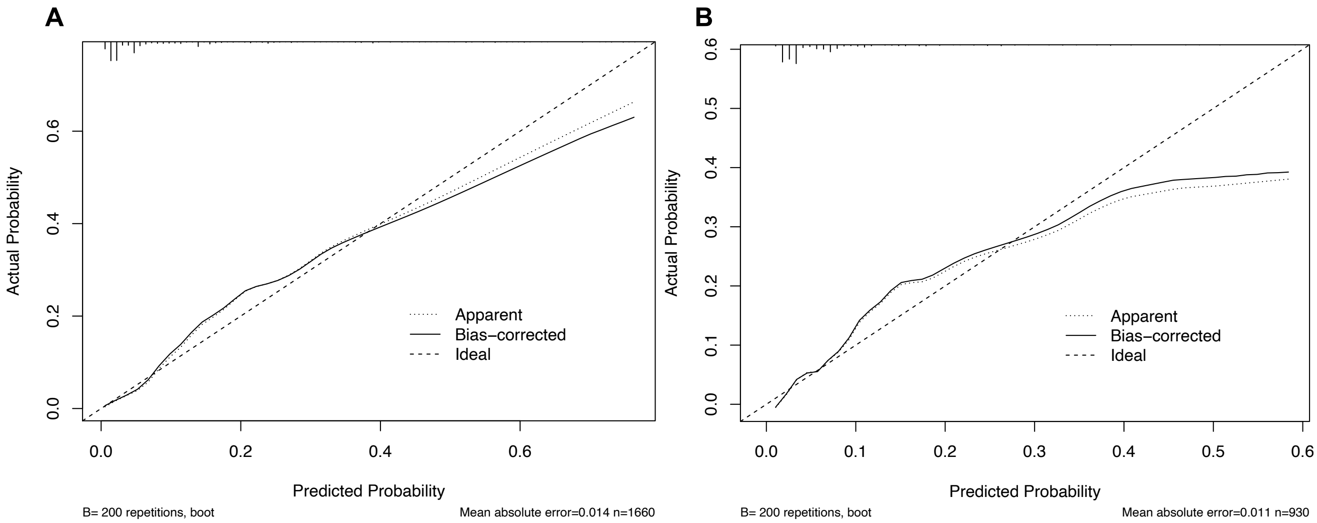


**Figure S2.** Calibration curves of the developed score predicting diabetes in the development cohort (A) and the external validation cohort (B).

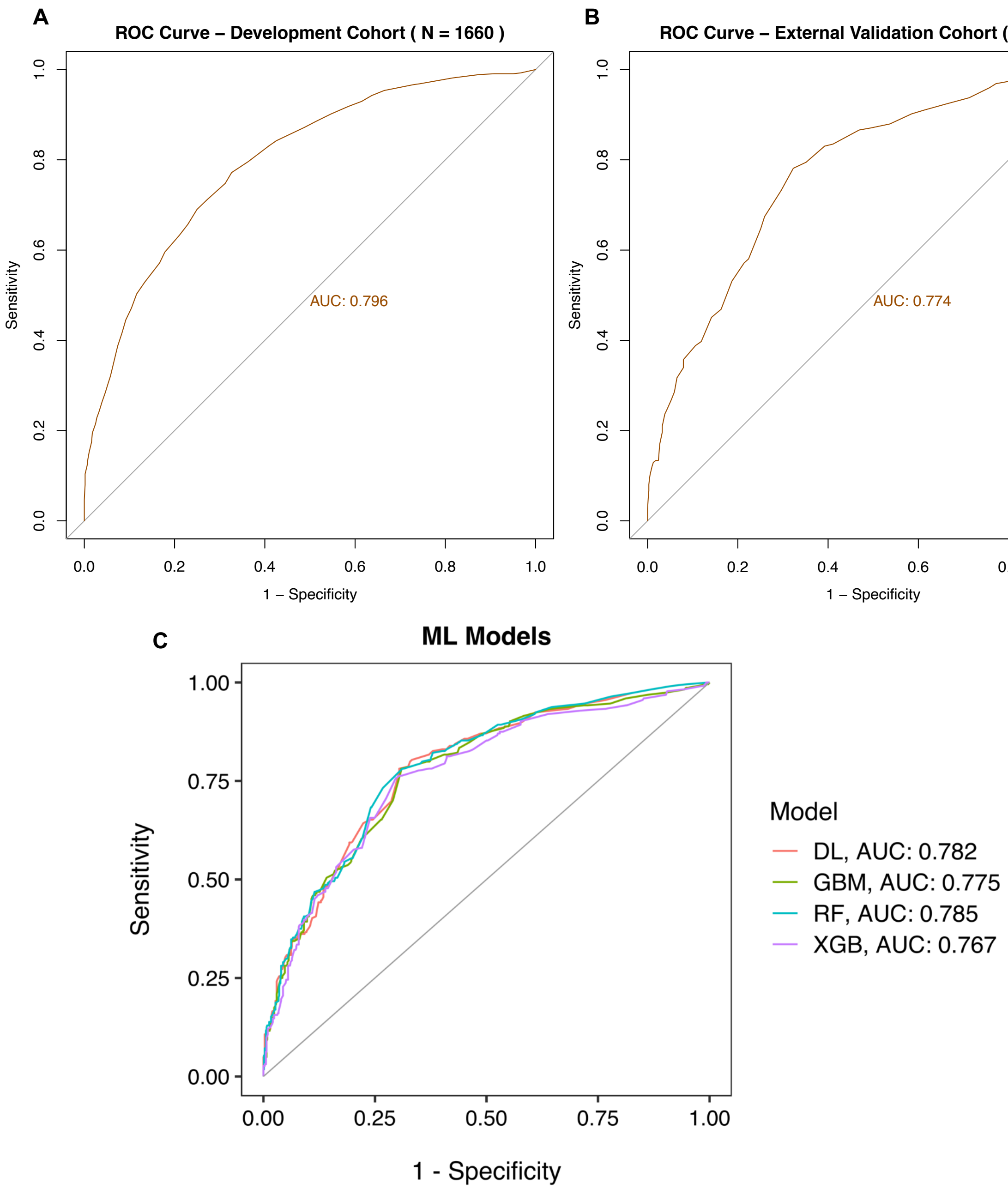


**Figure S3.** ROC curve showing the performance of developed risk score in predicting Impaired Glucose Metabolism (IGM) with AUC, 0.796 (95% CI: 0.774-0.819) in the development cohort (A) and 0.774 (95% CI: 0.740-0.809) in the external validation cohort (B). ROC curves for predicting Impaired Glucose Metabolism (IGM) using machine learning models. Deep learning (DL), gradient boosting machine (GBM), random forest (RF), XgBoost (XGB) (C).

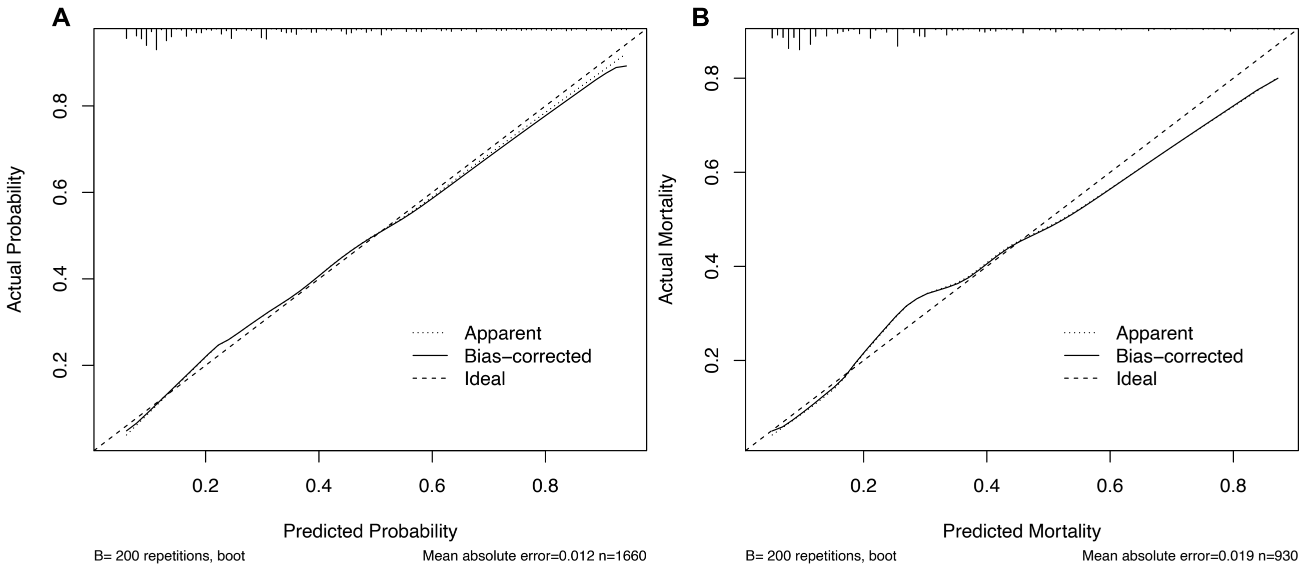


**Figure S4.** Calibration curves of the developed score predicting Impaired Glucose Metabolism (IGM) in the training cohort (A) and the validation cohort (B).

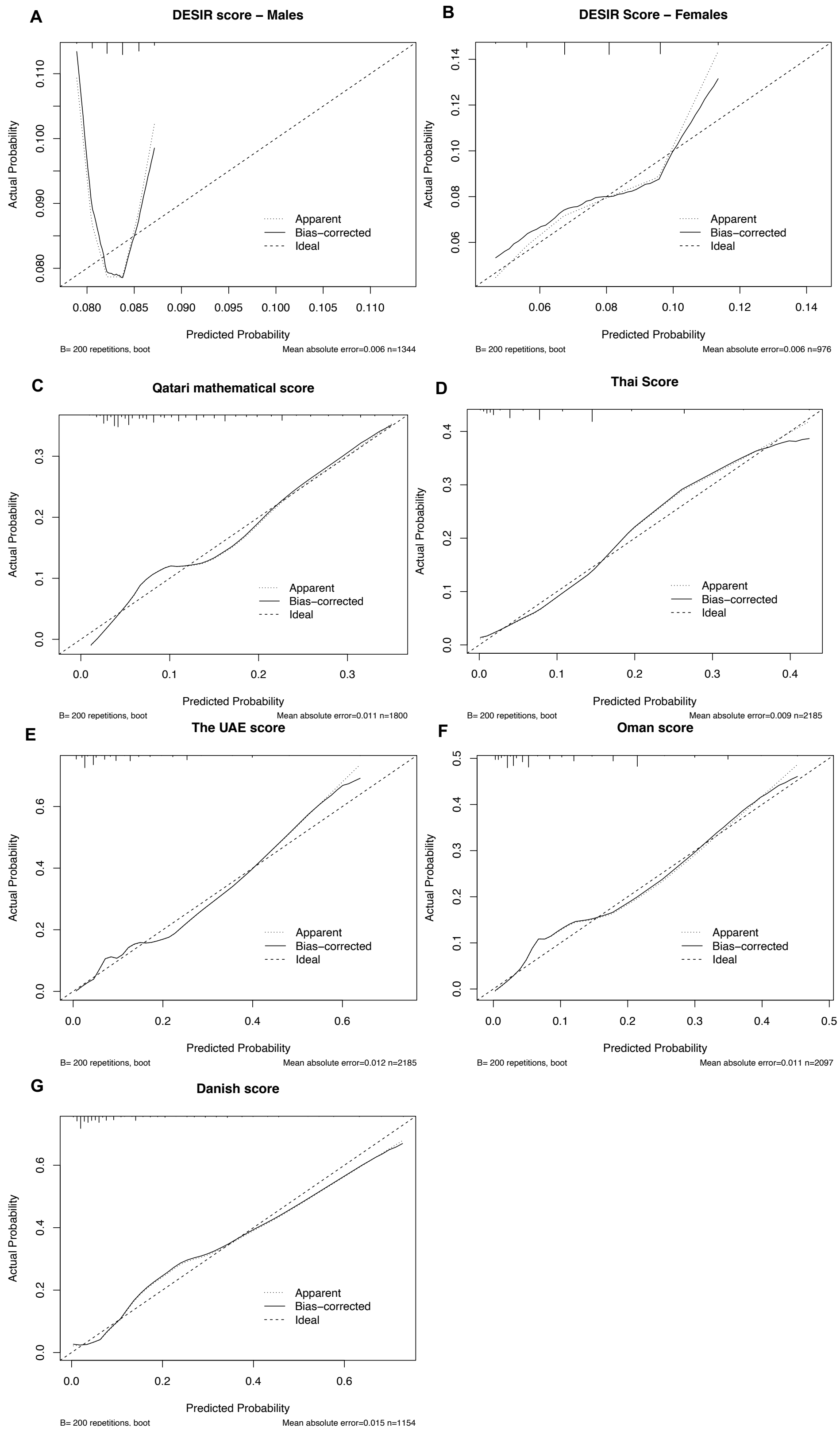


**Figure S5.** Calibration curves of the seven externally validated risk scores showing comparable performance except for the DESIR scores (A & B).

**Table S1**: Area under the curve (AUC), sensitivity, specificity, positive predictive value (PPV), negative predictive value (NPP) of the developed risk score values in the development and external validation cohorts.

| Cut-off | Specificity % | Sensitivity % | PPV% | NPV % |
|---|---|---|---|---|
| **Development Cohort** | | | | |
| 1 | .34 | 100 | 10.21 | 100 |
| 2 | 3.55 | 100 | 10.52 | 100 |
| 3 | 10.73 | 99.41 | 11.21 | 99.38 |
| 4 | 12.68 | 99.41 | 11.43 | 99.47 |
| 7 | 16.83 | 99.41 | 11.93 | 99.6 |
| 8 | 16.9 | 99.41 | 11.94 | 99.6 |
| 9 | 17.17 | 99.41 | 11.97 | 99.61 |
| 10 | 18.04 | 99.41 | 12.09 | 99.63 |
| 11 | 22.27 | 98.82 | 12.59 | 99.4 |
| 12 | 31.59 | 98.22 | 14 | 99.37 |
| 13 | 34.54 | 97.04 | 14.39 | 99.04 |
| 14 | 41.99 | 96.45 | 15.86 | 99.05 |
| 15 | 48.42 | 95.86 | 17.4 | 99.04 |
| 16 | 49.23 | 95.86 | 17.63 | 99.06 |
| 17 | 51.24 | 95.86 | 18.22 | 99.09 |
| 18 | 51.78 | 95.86 | 18.39 | 99.1 |
| 19 | 52.58 | 95.86 | 18.64 | 99.12 |
| 20 | 53.12 | 95.27 | 18.72 | 99 |
| 21 | 54.19 | 94.67 | 18.98 | 98.9 |
| 22 | 55.87 | 94.08 | 19.46 | 98.81 |
| 23 | 60.76 | 91.12 | 20.84 | 98.37 |
| 24 | 67.81 | 88.76 | 23.81 | 98.16 |
| 25 | 69.42 | 88.76 | 24.75 | 98.2 |
| 26 | 73.04 | 86.98 | 26.78 | 98.02 |
| 27 | 75.25 | 85.21 | 28.07 | 97.82 |
| 28 | 75.86 | 85.21 | 28.57 | 97.84 |
| 29 | 77.13 | 84.02 | 29.4 | 97.71 |
| 30 | 77.87 | 83.43 | 29.94 | 97.65 |
| 31 | 79.48 | 82.25 | 31.24 | 97.53 |
| 32 | 80.48 | 79.88 | 31.69 | 97.24 |
| 33 | 81.22 | 78.11 | 32.04 | 97.04 |
| 34 | 82.56 | 76.92 | 33.33 | 96.93 |
| 35 | 84.37 | 73.37 | 34.73 | 96.55 |
| 36 | 87.32 | 67.46 | 37.62 | 95.95 |
| 37 | 88.46 | 64.5 | 38.79 | 95.65 |
| 38 | 90.01 | 59.76 | 40.4 | 95.18 |
| 39 | 90.27 | 57.99 | 40.33 | 94.99 |
| 40 | 90.74 | 57.4 | 41.28 | 94.95 |
| 41 | 91.62 | 55.03 | 42.66 | 94.73 |
| 42 | 92.29 | 52.07 | 43.35 | 94.44 |

| | | | | |
|---|---|---|---|---|
| 43 | 93.83 | 47.34 | 46.51 | 94.02 |
| 44 | 94.43 | 46.15 | 48.45 | 93.93 |
| 45 | 94.84 | 43.2 | 48.67 | 93.64 |
| 46 | 95.04 | 42.6 | 49.32 | 93.59 |
| 47 | 95.31 | 42.01 | 50.35 | 93.55 |
| 48 | 95.51 | 39.05 | 49.62 | 93.25 |
| 49 | 96.24 | 34.32 | 50.88 | 92.82 |
| 50 | 97.52 | 29.59 | 57.47 | 92.43 |
| 51 | 97.65 | 27.22 | 56.79 | 92.21 |
| 52 | 97.79 | 26.04 | 57.14 | 92.1 |
| 54 | 97.85 | 24.85 | 56.76 | 91.99 |
| 55 | 98.05 | 22.49 | 56.72 | 91.78 |
| 56 | 98.32 | 14.79 | 50 | 91.06 |
| 57 | 98.66 | 10.65 | 47.37 | 90.69 |
| 58 | 98.99 | 9.47 | 51.61 | 90.61 |
| 59 | 99.2 | 8.28 | 53.85 | 90.51 |
| 61 | 99.26 | 8.28 | 56 | 90.52 |
| 62 | 99.33 | 6.51 | 52.38 | 90.36 |
| 63 | 99.6 | 3.55 | 50 | 90.11 |
| 64 | 99.66 | 2.37 | 44.44 | 90.01 |
| 65 | 99.93 | 0.59 | 50 | 89.87 |
| **External Validation Cohort** | | | | |
| 1 | 1.38 | 100 | 6.64 | 100 |
| 2 | 4.49 | 100 | 6.85 | 100 |
| 3 | 12.66 | 100 | 7.44 | 100 |
| 4 | 15.19 | 100 | 7.64 | 100 |
| 8 | 21.06 | 100 | 8.17 | 100 |
| 9 | 21.4 | 100 | 8.2 | 100 |
| 10 | 23.59 | 100 | 8.41 | 100 |
| 11 | 27.62 | 98.36 | 8.71 | 99.59 |
| 12 | 37.17 | 96.72 | 9.75 | 99.38 |
| 13 | 42.69 | 95.08 | 10.43 | 99.2 |
| 14 | 53.74 | 91.8 | 12.23 | 98.94 |
| 15 | 57.88 | 85.25 | 12.44 | 98.24 |
| 16 | 58.92 | 85.25 | 12.71 | 98.27 |
| 17 | 60.41 | 85.25 | 13.13 | 98.31 |
| 18 | 61.1 | 85.25 | 13.33 | 98.33 |
| 19 | 61.91 | 85.25 | 13.58 | 98.35 |
| 20 | 63.18 | 85.25 | 13.98 | 98.39 |
| 21 | 64.1 | 80.33 | 13.57 | 97.89 |
| 22 | 66.86 | 72.13 | 13.25 | 97.16 |
| 23 | 71.69 | 70.49 | 14.88 | 97.19 |
| 24 | 78.02 | 68.85 | 18.03 | 97.27 |
| 25 | 79.52 | 67.21 | 18.72 | 97.19 |
| 26 | 83.66 | 65.57 | 21.98 | 97.19 |

| | | | | |
|---|---|---|---|---|
| 27 | 84.23 | 63.93 | 22.16 | 97.08 |
| 28 | 84.7 | 60.66 | 21.76 | 96.84 |
| 29 | 85.62 | 59.02 | 22.36 | 96.75 |
| 30 | 86.88 | 54.1 | 22.45 | 96.42 |
| 31 | 89.3 | 50.82 | 25 | 96.28 |
| 32 | 90.33 | 49.18 | 26.32 | 96.2 |
| 33 | 90.79 | 49.18 | 27.27 | 96.22 |
| 34 | 91.14 | 45.9 | 26.67 | 96 |
| 35 | 91.83 | 40.98 | 26.04 | 95.68 |
| 36 | 92.75 | 37.7 | 26.74 | 95.5 |
| 37 | 93.56 | 32.79 | 26.32 | 95.2 |
| 38 | 94.71 | 31.15 | 29.23 | 95.14 |
| 39 | 95.28 | 29.51 | 30.51 | 95.06 |
| 40 | 95.28 | 27.87 | 29.31 | 94.95 |
| 41 | 95.28 | 26.23 | 28.07 | 94.85 |
| 42 | 95.63 | 24.59 | 28.3 | 94.75 |
| 43 | 96.32 | 21.31 | 28.89 | 94.58 |
| 44 | 97.35 | 19.67 | 34.29 | 94.53 |
| 45 | 97.7 | 18.03 | 35.48 | 94.44 |
| 46 | 97.81 | 18.03 | 36.67 | 94.44 |
| 47 | 97.81 | 14.75 | 32.14 | 94.24 |
| 48 | 98.16 | 13.11 | 33.33 | 94.15 |
| 49 | 98.27 | 11.48 | 31.82 | 94.05 |
| 50 | 98.62 | 11.48 | 36.84 | 94.07 |
| 51 | 98.96 | 6.56 | 30.77 | 93.78 |
| 54 | 99.31 | 6.56 | 40 | 93.8 |
| 55 | 99.42 | 4.92 | 37.5 | 93.71 |
| 56 | 99.54 | 4.92 | 42.86 | 93.72 |
| 60 | 99.65 | 3.28 | 40 | 93.62 |
| 62 | 99.65 | 1.64 | 25 | 93.52 |
| 63 | 99.65 | 0 | 0 | 93.42 |
| 64 | 99.77 | 0 | 0 | 93.43 |
| 71 | 99.88 | 0 | 0 | 93.43 |

*Abbreviations: PPV, positive predictive value; NPV, negative predictive value.*

**Table S2**. Aaccuracy (ACC), specificity and sensitivity of the risk score values in the machine learning algorithms predicting T2DM.

| Algorithm | Probability Cutoff | Training | | | |
|---|---|---|---|---|---|
| | | ACC (%) | Specificity (%) | Sensitivity (%) | AUC (%) |
| Deep Learning | .25 | 86.39 | 89.13 | 62.13 | .874 |
| | .5 | 89.52 | 98.46 | 10.65 | |
| | .75 | 89.70 | 99.80 | 0.59 | |
| Gradient boosting machine | .25 | 88.55 | 91.15 | 65.68 | .900 |
| | .5 | 91.51 | 98.32 | 31.36 | |
| | .75 | 90.06 | 99.93 | 2.96 | |
| Random Forest | .25 | 87.05 | 89.20 | 68.05 | .879 |
| | .5 | 90.66 | 99.33 | 14.20 | |
| | .75 | 90.06 | 99.73 | 4.73 | |
| XGBoost | .25 | 88.73 | 90.95 | 69.23 | .906 |
| | .5 | 91.87 | 98.32 | 34.91 | |
| | .75 | 90.54 | 99.80 | 8.88 | |
| **Algorithm** | **Probability Cutoff** | **External Validation** | | | |
| | | ACC (%) | Specificity (%) | Sensitivity (%) | AUC |
| Deep Learning | .25 | 90.11 | 93.44 | 42.62 | .817 |
| | .5 | 93.55 | 99.65 | 6.56 | |
| | .75 | 93.33 | 99.88 | 0.00 | |
| Gradient boosting machine | .25 | 89.68 | 93.44 | 36.07 | .745 |
| | .5 | 92.26 | 98.27 | 6.56 | |
| | .75 | 93.33 | 99.88 | 0.00 | |
| Random Forest | .25 | 89.57 | 93.10 | 39.34 | .813 |
| | .5 | 93.12 | 99.42 | 3.28 | |
| | .75 | 93.66 | 100.00 | 3.28 | |
| XGBoost | .25 | 89.14 | 93.79 | 22.95 | .717 |
| | .5 | 92.37 | 98.04 | 11.48 | |
| | .75 | 93.23 | 99.77 | 0.00 | |

**Table S3**: Multivariable logistic regression and assigned scores for variable predicting Impaired Glucose Metabolism (IGM).

| Variable | β coefficient | P value | OR (95% CI) | Risk score † |
|---|---|---|---|---|
| **Age (years)** | | | | |
| 18–36 | Reference | | | |
| 36–54 | 1.22 | <.001 | 3.39 (2.56–4.51) | 12 |
| ≥55 | 2.37 | <.001 | 10.76 (7.03–16.69) | 24 |
| **Gender** | | | | |
| Female | Reference | | | |
| Male | .27 | .02 | 1.32 (1.03–1.68) | 3 |
| **Waist-to-hip-ratio** | | | | |
| Normal | Reference | | | |
| Abnormal | .68 | <.001 | 1.98 (1.34–2.92) | 7 |
| **BMI (kg/m$^2$)** | | | | |
| Normal | Reference | | | |
| Overweight | .39 | .02 | 1.48 (1.05–2.10) | 4 |
| Obese | .64 | <.001 | 1.89 (1.34–2.69) | 6 |
| **History of hyperlipidemia(yes)** | .63 | <.001 | 1.88 (1.45–2.42) | 6 |
| **History of hypertension (yes)** | .51 | .002 | 1.67 (1.20–2.31) | 5 |
| **Education** | | | | |
| University/Postgraduate | Reference | | | |
| High-school or less than university | .16 | .22 | 1.18 (.89–1.55) | 2 |
| Secondary school – absent school education | .69 | .001 | 2.01 (1.32–3.07) | 7 |
| **Vegetable consumption (< once daily)** | - | - | - | 1 |
| **Fruit consumption (< once daily)** | - | - | - | 1 |
| **Cutoff point †** | | | | **25** |

*Abbreviations: OR, odds ratio; CI, confidence interval.*
† The cut-off point was obtained from the optimal Youden index.
values 0-25, 26-36, >36 were deemed as low, moderate, and high risk of developing Impaired Glucose Metabolism (IGM, respectively).

**Table S4**: Scores allocated to variables used in the externally validated risk scores.

| Model / Variable | β coefficient | OR | OR (95% CI) | Risk score |
|---|---|---|---|---|
| **The Danish model [1]** | | | | |
| **Age** | | | | |
| 45 | .6926 | 2.0 | (1.0 - 4.1) | 7 |
| 50 | 1.3111 | 3.7 | (2.0 - 7.0) | 13 |
| 55-60 | 1.8475 | 6.3 | (3.5 - 11.5) | 18 |
| **Male gender** | .3970 | 1.5 | (1.0 - 2.2) | 4 |
| **BMI** | | | | |
| 25-29 | .7401 | 2.1 | (1.3 - 3.5) | 7 |
| 30 | 1.4672 | 4.4 | (2.6 - 7.3) | 15 |
| **Known hypertension (yes)** | .9832 | 2.7 | (1.8 - 4.0) | 10 |
| **Physically inactive** | .6488 | 1.9 | (1.0 - 3.5) | 6 |
| **Parent having diabetes (yes)** | .6835 | 2.0 | (1.3 - 3.0) | 7 |
| **The Omani model [2]** | | | | |
| **Age** | | | | |
| 40-59 | 1.8 | 5.9 | (4.5 - 7.7) | 7 |
| ≥ 60 | 2.3 | 9.7 | (6.9 - 13.8) | 9 |
| **Waist circumference (Men ≥94 cm, women ≥80 cm)** | | | | |
| **BMI (kg/m$^2$)** | | | | |
| 25 to <30 | .54 | 1.7 | (1.3 - 2.3) | 2 |
| ≥30 | .69 | 2.0 | (1.4 - 2.8) | 3 |
| **Family history of diabetes (yes)** | 1.9 | 6.9 | (5.4 - 8.8) | 8 |
| **Current hypertension status (yes)** | .73 | 2.1 | (1.6 - 2.6) | 3 |
| **The UAE model [3]** | | | | |
| **Age** | | | | |
| 35-64 | .424 | 1.75 | (1.10 - 3.38) | 4 |
| ≥ 65 | 1.017 | 3.38 | (2.23 - 5.12) | 10 |
| **Family history of diabetes (parent/siblings)** | .734 | 2.08 | (1.47 - 2.96) | 7 |
| **Hypertension status (yes)** | .389 | 1.48 | (1.04 - 2.18) | 4 |
| **BMI (kg/m$^2$)** | | | | |
| ≥ 30.0 | .676 | 1.97 | (1.17 - 3.30) | 7 |
| **Waist-to-hip ratio** (Men ≥0.90, women ≥0.85) | .534 | 1.71 | (1.08 - 2.69) | 5 |
| **The DESIR model – Males [4]** | | | | |
| **Waist circumference (cm)** | | | | |
| 80-89 | | | | 1 |
| 90-99 | | | | 2 |
| ≥ 100 | | | | 3 |
| **Current smoker (yes)** | | | | 1 |
| **Hypertension (yes)** | | | | 1 |

| | | | | |
|---|---|---|---|---|
| **The DESIR models – Females [4]** | | | | |
| **Waist circumference (cm)** | | | | |
| 70-79 | | | | 1 |
| 80-89 | | | | 2 |
| ≥ 90 | | | | 3 |
| **Diabetes in family (yes)** | | | | 1 |
| **Hypertension (yes)** | | | | 1 |
| **The Thai model [5]** | | | | |
| **Age (years)** | | | | |
| 40–44 | −.07 | | | 0 |
| 45–49 | .27 | | | 1 |
| ≥50 | .60 | | | 2 |
| **Male gender** | .44 | | | 2 |
| **BMI (kg/m2)** | | | | |
| ≥23 - <27.5 | .69 | | | 3 |
| ≥27.5 | 1.24 | | | 5 |
| **Waist circumference (≥90 cm in men, ≥80 cm in women)** | .56 | | | 2 |
| **Hypertension (yes)** | .64 | | | 2 |
| **History of diabetes in parent/sibling (yes)** | 1.08 | | | 4 |
| **The Qatari mathematical model [6]** | | | | |
| **Age (years)** | | | | |
| 20-24 | 59 | 2.24 | (1.39 - 3.62) | 6 |
| 25-29 | 1.02 | 4.01 | (2.55 - 6.32) | 10 |
| 30-34 | 0.97 | 4.63 | (2.94 - 7.28) | 10 |
| 35-39 | 1.42 | 7.65 | (4.92 - 11.89) | 14 |
| 40-44 | 1.54 | 9.16 | (5.84 - 14.37) | 15 |
| 45-49 | 1.79 | 11.82 | (7.56 - 18.48) | 18 |
| 50-54 | 2.02 | 13.08 | (8.31 - 20.60) | 20 |
| 55-59 | 2.46 | 20.48 | (13.03 - 32.20) | 25 |
| 60-64 | 2.46 | 18.00 | (11.23 - 28.85) | 25 |
| 65-69 | 2.39 | 15.40 | (9.41 - 25.23) | 24 |
| 70-74 | 2.62 | 15.75 | (9.30 - 26.68) | 26 |
| 75-79 | 2.45 | 13.57 | (7.55 - 24.39) | 24 |
| **Male gender** | .20 | 1.13 | (0.98 - 1.3) | 2 |
| BMI ≥ 30.0 | 1.40 | 4.53 | (3.89 - 5.28) | 14 |
| Smoker | .31 | 1.5 | (1.26 - 1.79) | 3 |
| **Physically** Inactive | .53 | 2.26 | (1.95 - 2.61) | 5 |

**Table S5**: Comparison of validation metrics between the model developed (Qatari score) and externally validated models in predicting Impaired Glucose Metabolism (IGM) and Type two Diabetes Mellites (T2DM).

| **Metric[a]** | **N** | **AUC** | **McKelvoy's $R^2$** | **Brier score** | **Intercept** | **Calibration slope** | **HL $\chi^2$ test (df)** | **HL $\chi^2$ *P*-value** | **AIC** |
|---|---|---|---|---|---|---|---|---|---|
| **IGM** | | | | | | | | | |
| Qatari score | 930 | .780 (.740 - .80) | .262 | .148 | -.003 | .9947 | 7.42 (8) | .492 | 848 |
| **T2DM** | | | | | | | | | |
| Qatari score (QDRISK) | 930 | .818 (.765 - .864) | .202 | .051 | .057 | 1.021 | 4.78 (8) | .780 | 381 |
| Danish score [1] | 1154 | .824 (.778 - .8694) | .281 | .069 | .020 | 1.004 | 13.65 (8) | .091 | 560 |
| UAE score [3] | 2185 | .781 (.7516 - .812) | .191 | .073 | .002 | 1.001 | 52.56 (6) | <.001 | 1127 |
| Omani score [2] | 2097 | .796 (.7673 - .8245) | .212 | .075 | .026 | 1.005 | 2.72 (7) | .004 | 1078 |
| Thai score [5] | 2185 | .766 (.7353 - .7999) | .170 | .074 | .007 | 1.003 | 13.07 (5) | .022 | 1151 |
| Qatari mathematical model [6] | 1800 | .733 (.697 - .770) | .124 | .081 | .025 | 1.011 | 15.08 (8) | .057 | 1034 |
| DESIR score – Females [4] | 976 | .555 (.508 - .635) | - | - | - | - | - | - | - |
| DESIR score – Males [4] | 1344 | .486 (.429 - .554) | - | - | - | - | - | - | - |

*Abbreviations:* AUC, area under the receiver operating characteristic curve; HL, Hosmer–Lemeshow goodness of fit test; *AIC, Akaike information criterion.*
[a] Values were generated using 200 bootstrap resamples.

### References


1. Glümer, C., et al., *A Danish diabetes risk score for targeted screening: the Inter99 study.* Diabetes Care, 2004. **27**(3): p. 727-33.
2. Al-Lawati, J.A. and J. Tuomilehto, *Diabetes risk score in Oman: a tool to identify prevalent type 2 diabetes among Arabs of the Middle East.* Diabetes Res Clin Pract, 2007. **77**(3): p. 438-44.
3. Sulaiman, N., et al., *Diabetes risk score in the United Arab Emirates: a screening tool for the early detection of type 2 diabetes mellitus.* BMJ Open Diabetes Res Care, 2018. **6**(1): p. e000489.
4. Balkau, B., et al., *Predicting diabetes: clinical, biological, and genetic approaches: data from the Epidemiological Study on the Insulin Resistance Syndrome (DESIR).* Diabetes Care, 2008. **31**(10): p. 2056-61.
5. Aekplakorn, W., et al., *A risk score for predicting incident diabetes in the Thai population.* Diabetes Care, 2006. **29**(8): p. 1872-7.
6. Awad, S.F., et al., *A diabetes risk score for Qatar utilizing a novel mathematical modeling approach to identify individuals at high risk for diabetes.* Sci Rep, 2021. **11**(1): p. 1811.
